# Uncovering the Post-Pandemic Timing of Influenza, RSV, and COVID-19 Driving Seasonal Influenza-like Illness in the United States

**DOI:** 10.1101/2025.08.21.25333432

**Authors:** George Dewey, Austin G. Meyer, Raul Garrido Garcia, Mauricio Santillana

**Affiliations:** Machine Intelligence Group for the betterment of Health and the Environment, Northeastern University, Boston, MA, USA, 02115; Network Science Institute, Northeastern University, Boston, MA 02115, USA; Department of Physics, Northeastern University, Boston, MA 02115, USA; Baylor Scott and White Health, Dallas, TX 75426, USA; Department of Epidemiology, Harvard T.H. Chan School of Public Health, Boston, MA, USA, 02115

**Keywords:** disease surveillance, RSV, influenza, influenza-like illness, forecasting, pandemic preparedness

## Abstract

**Background:** Influenza and respiratory syncytial virus (RSV) are major contributors to the burden of seasonal influenza-like illnesses (ILI) in the US. The prevention and treatment of ILI varies substantially across age groups and in cost and administration schedule. Clearly identifying the times when healthcare resources are most needed to mitigate the effects of seasonal RSV and influenza outbreaks will improve public health responses before and during ILI seasons.

**Methods:** We implemented stacked-regression linear models to infer the contribution of each of these diseases to seasonal ILI syndromic indicators. We further implemented anomaly-detection algorithms on data from the US Centers for Disease Control and Prevention National Syndromic Surveillance Program to identify the timing of onsets and peaks of RSV, influenza, and COVID-19.

**Findings:** A total of 148 state-ILI seasons were analyzed. In 114 out of 148 (77.0%) of analyzed seasons, volume of RSV emergency department (ED) visits peaked before influenza ED visits. The median time difference between peaks of RSV and peaks of influenza was +3.0 weeks. The timing of RSV and influenza onsets were found to occur more synchronously in the 2023-2024 and 2024-2025 ILI seasons.

**Interpretations:** RSV epidemics frequently reach peak volume before influenza epidemics across the US. Healthcare professionals and public health authorities should anticipate increases in RSV cases and hospitalizations at the start of the annual ILI season and establish infrastructure and planning to handle incoming surges of both RSV and influenza appropriately.

**Funding:** GD, RG, and MS were supported by cooperative agreement CDC-RFA-FT-23-0069 from the CDCs Center for Forecasting and Outbreak Analytics. AM was supported by the National Institutes of Health LRP #1L70AI194328-01. The views expressed in written in this publication do not necessarily reflect the official policies of the Department of Health and Human Services/Centers for Disease Control and Prevention.

**Summary:** Epidemics of respiratory pathogens such as influenza or RSV drive the influenza-like illness season in the US. We show that RSV epidemics peak before influenza epidemics in most states, with about a one to three week difference separating the epidemics.

## Introduction

Influenza-like illnesses (ILI), defined by the US Centers for Disease Control (CDC) as illnesses causing a fever (usually 100°F or 37.8°C or higher) plus cough and/or sore throat, affect more than 9 to 41 million people in the US every year^1^. ILI syndromic systems have operated for more than a decade in the United States (US)^2^. Indicators tracking ILI in the populations are frequently used as proxies of the timing of seasonal RSV and influenza outbreaks, which have historically started in autumn and ended by the beginning of spring. However, the COVID-19 pandemic brought about two major shifts to the respiratory disease landscape: first, it established the co-circulation of three pathogens with substantial epidemic potential which fall under the umbrella of ILI: influenza, respiratory syncytial virus (RSV), and SARS-CoV-2; second, it resulted in temporary enhancement of respiratory disease surveillance that enabled the disentanglement of these diseases during and between annual ILI seasons^3^.

While the symptom-based definition of ILI ensures that a majority of cases presenting to outpatient health facilities are detected by surveillance efforts, its lack of specificity results in the aggregation of cases of not only influenza A and B, RSV, and COVID-19, but also other respiratory infections before laboratory testing occurs^4^. Each of the component infections of infections demonstrate different levels of morbidity and mortality, especially across age groups; for example, RSV infection is associated with higher rates of lower respiratory infection in young children^5^ while influenza, RSV, and COVID-19 are all associated with increased mortality among people aged 65 and over^6,7^. Additionally, antiviral treatments for severe cases of these infections differ, with no overlap between US-approved treatments for the different viruses^8-10^. Misidentification or misconceptions about the infections aggregated into ILI has substantial consequences for public health response and outbreak preparedness: although many ILI cases in the US are not life-threatening, surges in the volume of respiratory infections can stress health infrastructure and medical providers, especially as care regimens for different infections differ in cost and complexity.

Because respiratory illness surveillance, prevention, and interventions have historically focused on influenza in the US^11-13^, healthcare systems and personnel are much more optimized to combat influenza outbreaks compared to outbreaks of other diseases. For example, while an influenza vaccination for a child age 6 months and older costs between US $15 and US $50 for a single dose, a comparable RSV vaccination costs between US $250 and US $500 per dose^14^. Furthermore, the public health focus on influenza has also led to the persistence of misconceptions related to ILI and influenza among healthcare workers, such as believing peaks in ILI activity are caused only by outbreaks of influenza A followed by outbreaks of influenza B^15-17^. Fortunately, new data streams introduced during the COVID-19 pandemic show promise in the ability to clearly distinguish between trends of influenza, RSV, and COVID-19 and to predict the onset and peaks of epidemics of these infections^18^.

Anticipating the timing of respiratory epidemics that drive the seasonal trajectory of ILI has several important implications for public health officials and medical practitioners. First, predicting the onset of an ILI season provides healthcare facilities and staff with advance notice to prepare for upcoming respiratory emergencies. Prediction algorithms and surveillance systems, such as those implemented by the CDC’s FluSight initiative^19,20^, aim at providing public health officials with valuable information to improve health communication and guide the allocation of resources, including both human personnel and protective equipment. Second, administration of therapeutics and vaccinations against respiratory infections is sensitive to timing, especially for at-risk populations such as young children or the elderly. For example, RSV prophylaxis for infants (a high-risk group for serious RSV infection) using the monoclonal antibody nirsevimab must be administered before the onset of an infant’s first RSV season; infants born during the RSV season should receive nirsevimab within one week of birth^21^. In addition, prior studies suggest passive immunity from nirsevimab wanes over time^22-24^, with the waning period matching the average duration of a single RSV season. As a result, it would be ideal to administer nirsevimab doses no earlier than necessary for any specific region of the US. Clearer understanding and prediction of the timing of the start of the ILI season, especially as it relates to the onset of seasonal RSV surges, would allow healthcare facilities to increase stock of needed treatments and brief concerned parents well before the seasonal surge in RSV cases occurs. Likewise, recent evidence suggests that immunity for influenza following vaccination undergoes significant waning after a few months^25,26^. Therefore, timing for influenza vaccination and RSV experience similar public health constraints and distinguishing the two during an ILI season is critical to public health response.

With these benefits in mind, this study aimed to improve our understanding of the observed timing and ordering of respiratory epidemics in the US that contribute to the trajectory of seasonal ILI. Using state-level surveillance data on emergency department (ED) visits from the CDC’s National Syndromic Surveillance System (NSSP)^27^ from the 2022-23, 2023-24, and 2024-25 ILI seasons, we show that the peak volume of RSV emergency department visits occurred before the peak volume of influenza emergency department visits in 77% of the analyzed state-ILI seasons, with a median difference of 3 weeks (range: -8, +10 weeks) between peaks of RSV epidemics and the peaks of influenza epidemics. We additionally highlight the co-circulation and irregular seasonality of COVID-19 during the evaluated ILI seasons. Our results indicate that RSV epidemics initiate most ILI seasons and that RSV epidemics are generally followed by epidemics of influenza.

## Methods

### Data sources

This study utilizes publicly available data collected by two major CDC surveillance systems: the US Outpatient influenza-like Illness Surveillance Network (ILINet) and the National Syndromic Surveillance Program (NSSP). ILINet is a collaboration between national, state, and local health departments and healthcare facilities that records the volume of hospital visits attributed to any influenza-like illness and submits those records to the CDC for centralized data storage. Reports are made based on patient visits due to ILI, where ILI is defined (using the World Health Organization definition) as “fever” (temperature of 100°F [37.8°C] or greater) and a cough and/or a sore throat”. NSSP records the percentage of US emergency department visits attributable to influenza, COVID-19, or RSV. In contrast with respiratory surveillance systems with longer data collection periods (such as CDC’s RSV-NET), data from 49 states and the District of Columbia has been stored by NSSP (data from Missouri is not available) since mid-2022.

The final analysis incorporated 148 state-ILI seasons (out of a possible 153) spanning the 22-23, 23-24, and 24-25 ILI seasons. Three state-ILI seasons from Missouri were excluded because the NSSP dataset does not include any data from that state. Similarly, we excluded the 24-25 seasons for Wyoming and Vermont from the analysis due to data unavailability; Wyoming supplied only data on ILI, while the NSSP time series from Vermont was available only up to data from September 28, 2024. Analysis of the 22-23 ILI season does not include the entire seasonal date range, as the first available data from NSSP is from October 1, 2022. All statistical analysis was performed between January 2025 and April 2025, with the last included date for both ILINet and NSSP data being March 30, 2025.

### Estimating the contribution of influenza, RSV and COVID-19 signals to ILI indicators

We used a stacked regression scheme^28^ to evaluate the contributions of the volume of individual respiratory outbreak signals to the trajectory of seasonal ILI and to determine the ordering of influenza, RSV, and COVID-19 outbreaks across the study period. We first filtered the national data to generate individual datasets for each state-ILI season, defining a state-ILI season as beginning in May of one year and ending in May of the following year. We use this definition to analyze data from a complete year, as traditional influenza surveillance records typically exclude the summer months (i.e., June, July, and August) when influenza cases are rare. Next, we rescaled each signal to constrain signal values to the range [0, 1], where a value of 1 represents the maximum signal strength and 0 represents the minimum signal strength. We then construct ridge regression models, using a grid-search algorithm to obtain the best lambda values for each dataset. The regression formula for each state-season is:

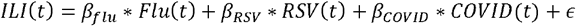

where ILI is the rescaled ILI volume for the timepoint *t* in each state and *ε* is a generic error term. For these regression models, we use the *penalized* R package ^29^ to remove the intercept and constrain predicted values for each individual predictor to be positive. This ensures that our processed signals have positive values for the entire study period. The result of this procedure is to generate sets of time series data which show the contribution of each individual respiratory illness to the overall volume of ILI for a single season. We then use the results of the modelling procedure to categorize the state-ILI seasons into seasons where peak RSV volume occurred before peak influenza volume (i.e., t_peak RSV_ < t_peak influenza_), seasons where peak RSV volume and peak influenza volume occurred in the same 7-day period (t_peak RSV_ = t_peak influenza_), and seasons where peak RSV volume occurred after peak influenza volume (t_peak RSV_ > t_peak influenza_). In seasons with multiple peaks of any respiratory epidemic, we use the seasonal maximum volume value for RSV, influenza, and COVID-19 for comparison.

### Evaluating the time difference between epidemics

We also carried out analyses to determine the time difference between the peaks and onsets of RSV and influenza seasonal epidemic outbreaks. To perform these analyses, we used an anomaly detection methodology previously used by Kogan et al.^30^ and Stolerman et al.^31^ to determine the timing of the onsets of each epidemic time series. Specifically, for each week *t* of our time series, we assessed the preceding 6 weeks and determined if exponential growth of cases was observed. This was done by computing a retrospective linear regression model on the time series at week *t* to find the multiplicative constant that maps the number of cases at time *t* to those occurring at time *t+1* (i.e., the coefficient λ_*t*_ of a lag-1 autoregressive model without an intercept) using data from the immediate six-week time period preceding week *t*. λ_*t*_ is a proxy of the effective reproductive number R_t_ which is a commonly used epidemiological measure to determine whether an epidemic is growing or decreasing in size^30,31^. Outbreaks were identified as the first point in time when λ_*t*_ was larger than 1 for a sustained period of six weeks. We choose this six-week evaluation period to reduce the chance of mislabeling noisy oscillations in the data as outbreaks.

To determine epidemic peaks, we used the *find_peaks* function from the Python library SciPy to identify peak values of our disease time series. After identification, each peak was assessed and selected based on the following three criteria: 1) peaks had to span a minimum duration of 2.5 weeks; 2) the peak value had to exceed the 75th percentile of the historical time series for each state; and 3) each peak needed to be more than 20 weeks apart from any previously detected peak unless it was higher than a previously detected peak that had occurred within the last 20 weeks.

Given the criteria for peak and onset detection and the unavailability of data from before October 2022, we only used the EWS method to analyze the 23-24 and 24-25 seasons. For both methods, we calculated the median and 95% confidence intervals (CI) for the median time difference between two peaks or between two onsets for all assessed state-seasons. We obtained the CIs for the medians using the bootstrap method and 1,000 simulation iterations.

## Results

We analyzed 148 state-ILI seasons across the 2022-23, 2023-24, and 2024-25 ILI seasons (henceforth the 22-23, 23-24, and 24-25 seasons), encompassing data from 49 US states and the District of Columbia (**Figure 1**). From this data, we identified three main patterns of epidemic timings. In the most common and most relevant pattern, RSV epidemics reached peak volume before influenza epidemics (114/148 state-seasons, 77.0%). Secondary to this pattern were seasons in which RSV epidemics and influenza epidemics peaked concurrently (i.e., the peak volume of ED visits for both RSV and influenza occurred within the same 7-day period) (14/148 seasons, 9.5%) and seasons in which influenza epidemics peaked before RSV epidemics (20/148 seasons, 13.5%) (**Supplementary Figures 1-3**).

**Figure 1.**
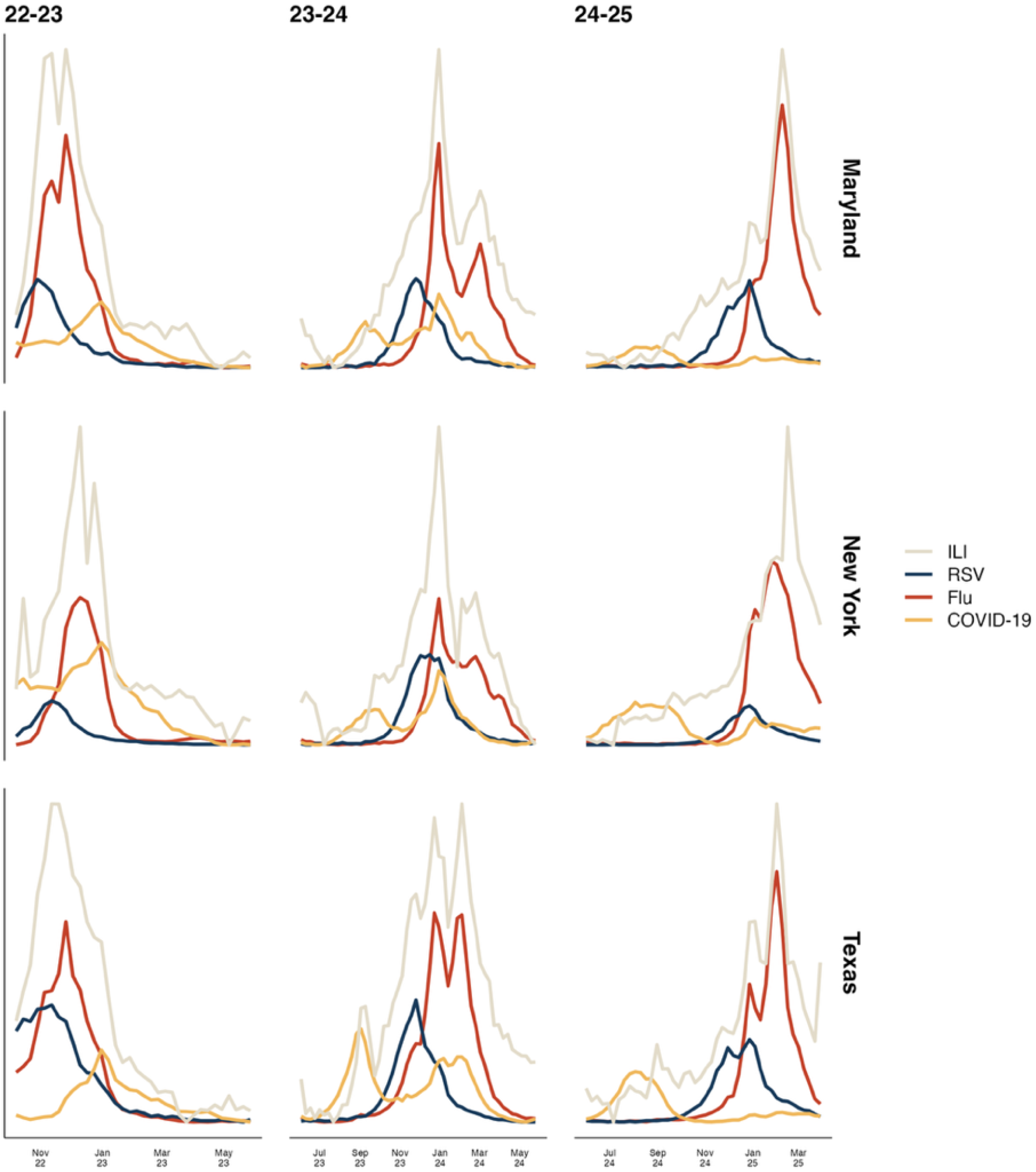
RSV epidemics precede influenza epidemics during many ILI seasons, while COVID-19 co-circulates. In each of the three states displayed (Maryland, New York, and Texas), RSV volume (blue) reaches its peak value before influenza volume (red) in each of the three analyzed seasons. COVID-19 volume (gold) peaked after both RSV and influenza in 22-23 but peaked before RSV and influenza in 23-24 and 24-25. Abbreviations - COVID-19: coronavirus disease 2019, flu: influenza, ILI: influenza-like illness, RSV: respiratory syncytial virus

ILI seasons most frequently reached their peak volume in November and December, with some late surges in February and March in the 23-24 and 24-25 seasons. Peak volume of RSV in the 22-23 season occurred as early as the first week of November (in Alabama, Mississippi, and Tennessee) and as late as the last week of December (in Montana). In the 23-24 season, the earliest RSV peak also occurred in the first week of November (in Florida); however, the latest peak occurred much later in the year, spiking in late February (in South Dakota). In the 24-25 season, the earliest RSV peak occurred in late November (in Florida, Georgia, and Mississippi) and the latest RSV peak occurred in early March (in Montana and South Dakota). Late peaks of ILI were observed in December (in the 22-23 season) and as late as March (in the 23-24 season). Across the three analyzed ILI seasons, COVID-19 did not demonstrate strictly annual seasonality like either RSV or influenza. While COVID-19 volume peaked between November and January in the 22-23 season, we observed multiple epidemics of COVID-19 in the 23-24 season in several states and only an early epidemic of COVID-19 in the 24-25 season (during the summer months of July – September).

We then used our early warning system (EWS) method to quantify the time differences between onsets and peaks of epidemics (**Figure 2**). First, we identified that the median difference between peaks of influenza and RSV epidemics was +2.0 weeks (95% percentile range [PR]: -5.0, +9.0) in the 23-24 season and +4.0 weeks (95% PR: -7.0, +6.2) in the 24-25 season (the + sign indicates that the RSV peak occurred before the influenza peak). Next, we also determined that the median time between the onsets of influenza and RSV epidemics was +3.0 weeks (95% PR: -7.4, +11.7) in the 23-24 season and +0.05 weeks in the 24-25 season (95% PR: -11.0, +11.0) (**Supplementary Figure 10**). The results of our analyses comparing peaks and onsets of COVID-19 to peaks and onsets of influenza were inconclusive as the onset and peak pattern of COVID-19 in the 23-24 season differed substantially from the onset and peak pattern of COVID-19 in the 24-25 season. We observed two epidemics of COVID-19 in most states in the summer of 2023 and in the winter between 2023 and 2024 but only observed a summer surge in COVID-19 volume in the 2024-25 season. Because our comparison assumes an overall seasonal trajectory like that of ILI (i.e., regular, seasonal outbreaks occurring within a similar timeframe), the inconsistent seasonality of COVID-19 represents a challenge for future research.

**Figure 2.**
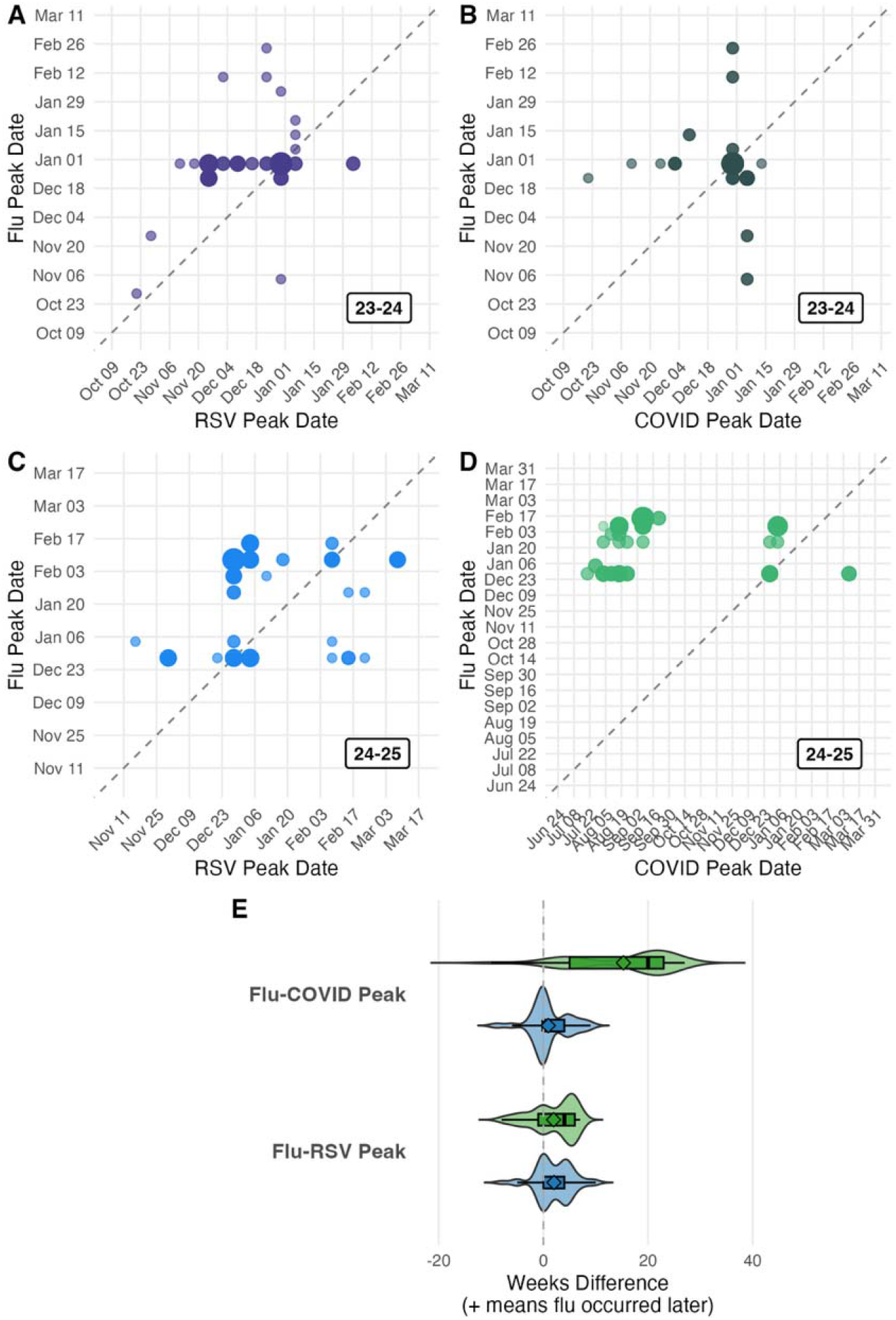
Early warning systems indicate RSV peaks before influenza in most seasons, while COVID-19 exhibits irregular seasonality. Points above the diagonal line indicate that the disease on the *y*-axis (influenza) peaks before the disease on the *x*-axis (either RSV or COVID-19). Larger points indicate that peaks in multiple states occurred during the same week. **A, B:** RSV peaked before influenza, while the COVID-19/influenza relationship is less clear in the 22-23 season. **C, D:** While the RSV-influenza relationship remains consistent, COVID-19 seasonality is noticeably different in the 24-25 season. **E:** Distribution and spread of the difference (in weeks) of RSV, COVID-19 and influenza peaks. Blue violins represent data from 23-24 and green violins represent data from 24-25.

## Discussion

In this study, we used data from the past three ILI seasons in the United States to clarify the temporal sequencing of epidemics of influenza and RSV. We deployed stacked regression models to evaluate the relative order of influenza, RSV and COVID-19 epidemics and their contribution to the overall trajectory of the ILI season. We determined that in 114 out of 148 recent state-ILI seasons (77.0%), RSV epidemics peaked before influenza epidemics. We then used an anomaly detection method to quantify the time differences between these epidemics and determined that there is a three-week median separation (range: -8, +10 weeks) between peaks of RSV epidemics and peaks of influenza epidemics. Taken together, these results suggest that public health authorities and healthcare providers should be ready to prepare for surges in RSV volume likely before surges in influenza volume (if this pattern persists), especially before and during the start of the traditional ILI season in early autumn.

The finding that the peaks of RSV epidemics preceded influenza epidemics in many of the recent ILI seasons in the United States has notable implications for public health authorities and healthcare providers. Based on our analyses, increases in the volume of RSV hospitalizations are a likely indicator for the start of the yearly ILI season. We also found that the ILI seasons we analyzed were consistently initiated by epidemics of RSV in the Southeastern United States; this geographic pattern aligns with the findings of previous work assessing trends in RSV^32^ in addition to work evaluating the spatiotemporal trends of other respiratory infections such as streptococcal pharyngitis^33^. The sources of this consistent outbreak initiation in usually warm Southeastern states during the start of winter should be investigated in future studies, especially as they potentially relate to changes in climate and in the environment. These findings also suggest that detection of surges of RSV cases in one state jurisdiction in late autumn or early winter should trigger alarms to health officials in nearby states for the onset of the annual ILI season, even before outpatient cases of RSV or influenza are reported.

Our findings also have direct implications for the administration of RSV prophylaxis with the monoclonal antibody nirsevimab. Current guidance from the Advisory Committee on Immunization Practices (ACIP) recommends seasonal administration from October through March for most of the continental United States ^34^. Moreover, the large number of children who primarily receive vaccines through the CDC’s Vaccines for Children (VFC) program means that state VFC programs generally cannot deviate from this window. However, our analyses reveal substantial state-level variability in the timing of RSV epidemic onsets and peaks. The broad, fixed window for nirsevimab administration is a practical compromise for national-level guidance, balancing the need to protect infants before the season starts against the realities of waning passive immunity and the significant cost of the therapeutic. A critical future direction is to use these data to quantify the portion of RSV cases that occur outside the standard ACIP window and to determine if these deviations follow predictable patterns. For instance, whether states in certain regions consistently experience later seasons would significantly impact nirsevimab administration policy through state VFC programs. Ultimately, our work suggests that a data-informed methodology to recommend administration windows on a state-by-state basis would be possible, allowing public health authorities to leverage real-time surveillance to issue more precise guidance and better align prophylaxis with the true local onset of the RSV season.

Additionally, while we focused on temporal relationships in this study, we recognize that the ordering, peaks, and onsets of respiratory epidemics may also be influenced by spatial factors. We observed a pattern of early-season epidemics occurring in the southeastern US that are followed by spreading across the country in a north-western direction for both influenza and RSV which resembled spatiotemporal patterns observed for other respiratory diseases as mentioned previously. We visualized this spread at the weekly level (**Supplementary Figures 11 and 12**) for both the 23-24 and 24-25 seasons and observed that the timings of RSV and influenza onsets both demonstrated the southeast to northwest spread pattern. On the other hand, peak timings for both influenza and RSV were more varied, with different sequences of peak timings occurring across the two analyzed seasons. While our interpretation of the spreading pattern across the country is limited by our weekly time resolution, we were still able to observe a gradual progression of epidemic onsets in a northwesterly direction. Future research should take advantage of finer-resolution data, such as data collected at the daily level, to track the movement of epidemics during ILI seasons.

While our results provide a top-down overview of the timing and ordering of peaks of RSV and influenza epidemics, questions about the reasons for the temporal separation between RSV and influenza remain. For example, while our findings concur with those of past studies evaluating the timing of RSV and influenza epidemics in Argentina^35^, other work done at the global scale^36^ identified that RSV epidemics in temperate regions (like the US) occurred before influenza epidemics, but that this relationship was not present in tropical regions. While this conclusion implicates weather and temperature changes as drivers of the relationship between RSV and influenza, it does not fully explain why RSV peaks before influenza in the US. One possible rationale for the earlier rise of RSV epidemics is that they are driven by the congregation of children of school age; however, since the average school opening occurs in late August to early September in the US^37^, it is unlikely that this congregation is the sole driver of RSV epidemics. It is therefore possible for a combination of climate and increased contact among younger individuals which promotes the growth of either RSV or influenza epidemics. Future work should carefully inspect drivers of respiratory epidemics through modeling or surveillance studies and identify the potential differences among those drivers.

Finally, this study has several limitations. First, our analyses relied on data reported to the CDC’s National Syndromic Surveillance System by participating emergency departments in hospitals nationwide and therefore do not necessarily capture the entire burden of ILI, influenza, or RSV cases. Second, our data were aggregated at the weekly level and therefore we could only temporally order individual epidemics within periods of 7 days. Lastly, we only had data for one fully complete ILI season and two mostly complete seasons due to the nature of the dataset used; future surveillance and analysis work should continue using this data moving forward to emphasize the robustness of our results.

## Acknowledgements

We thank Marc Lipsitch and William Hanage for helpful comments.

## Data Availability

Data and code used in this article can be found at github.com/MIGHTE-lab/rsv_peak_timing and will be released to the public upon article publication. The original data is provided by the United States Centers for Disease Control and Prevention through data.cdc.gov.

## Author Contributions

GD wrote the initial version of the manuscript and conducted the statistical analysis. GD, AM, and MS designed the study. GD, AM, RG, and MS reviewed the final version of the manuscript and approved it for submission.

## Figure Alt Text

Figure 1. Graphs showing the trajectories of seasonal ILI, influenza, RSV, and COVID-19 epidemics in a 3-by-3 grid for the 2022-23, 23-24, and 24-25 seasons.

Figure 2. Graphs and data about the relative timing of influenza, RSV, and COVID-19 peaks from early warning systems.

