## Supplementary Information for "Uncovering the Post-Pandemic Timing of Influenza, RSV, and COVID-19 Driving Seasonal Influenza-like Illness in the United States" (from another version of this preprint: version 1, 2025-08-26)

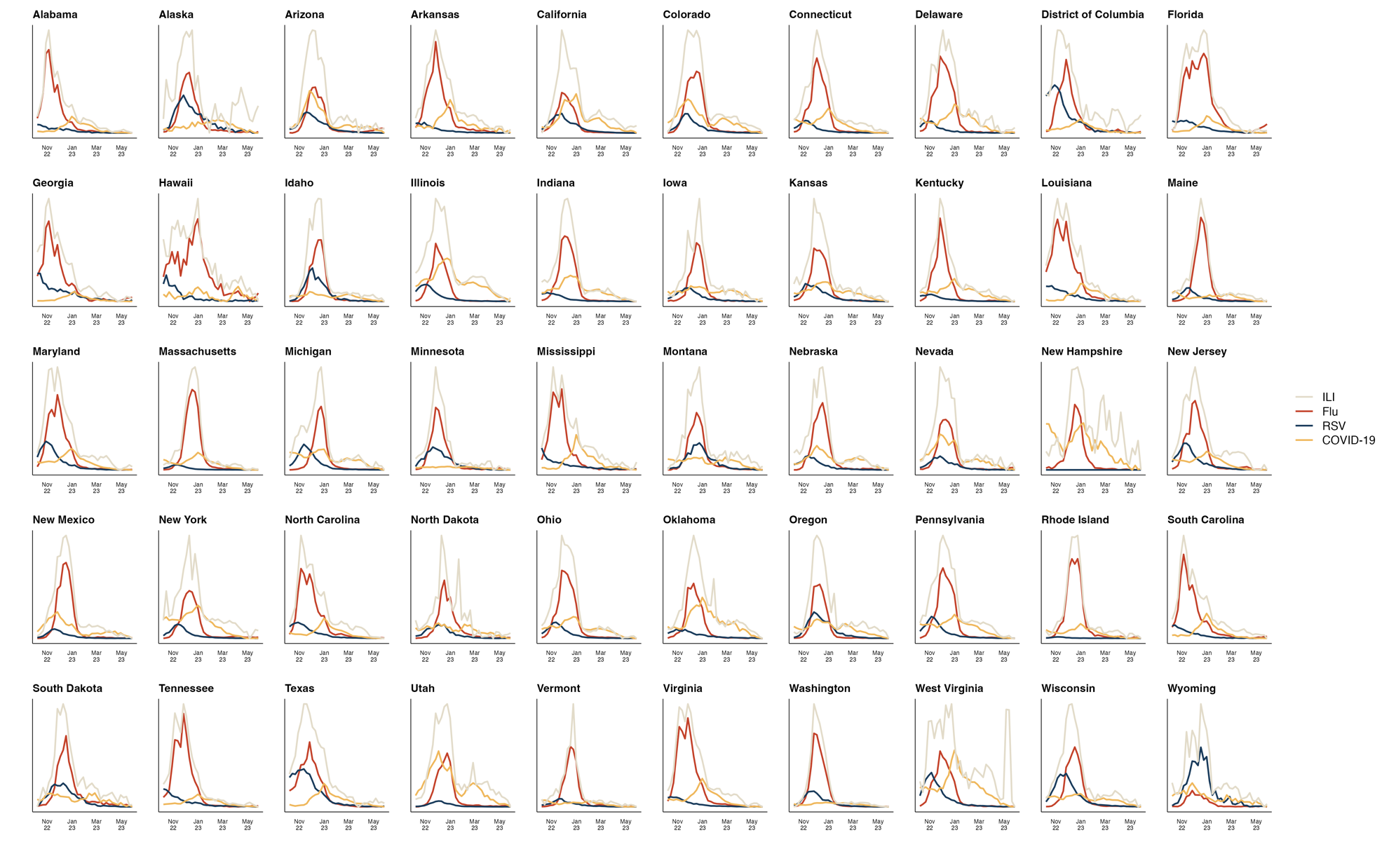

**Supplementary Figure 1: Timing of Respiratory Epidemics in the United States for the 2022-2023 ILI season.** Abbreviations - COVID-19: coronavirus disease 2019; flu: influenza; ILI: influenza-like illness; RSV: respiratory syncytial virus

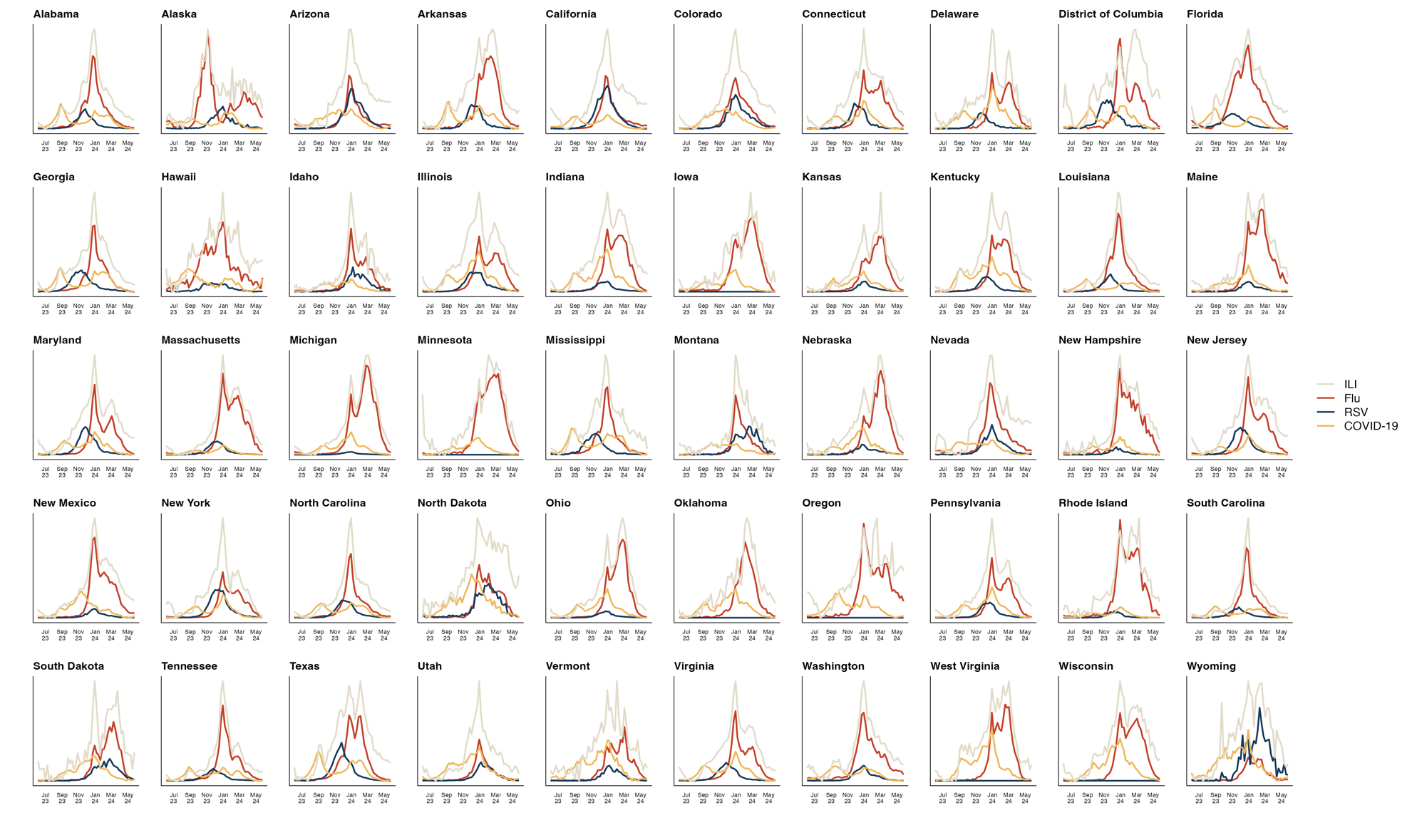

**Supplementary Figure 2: Timing of Respiratory Epidemics in the United States for the 2023-2024 ILI season.** Abbreviations - COVID-19: coronavirus disease 2019; fFu: influenza; ILI: influenza-like illness; RSV: respiratory syncytial virus

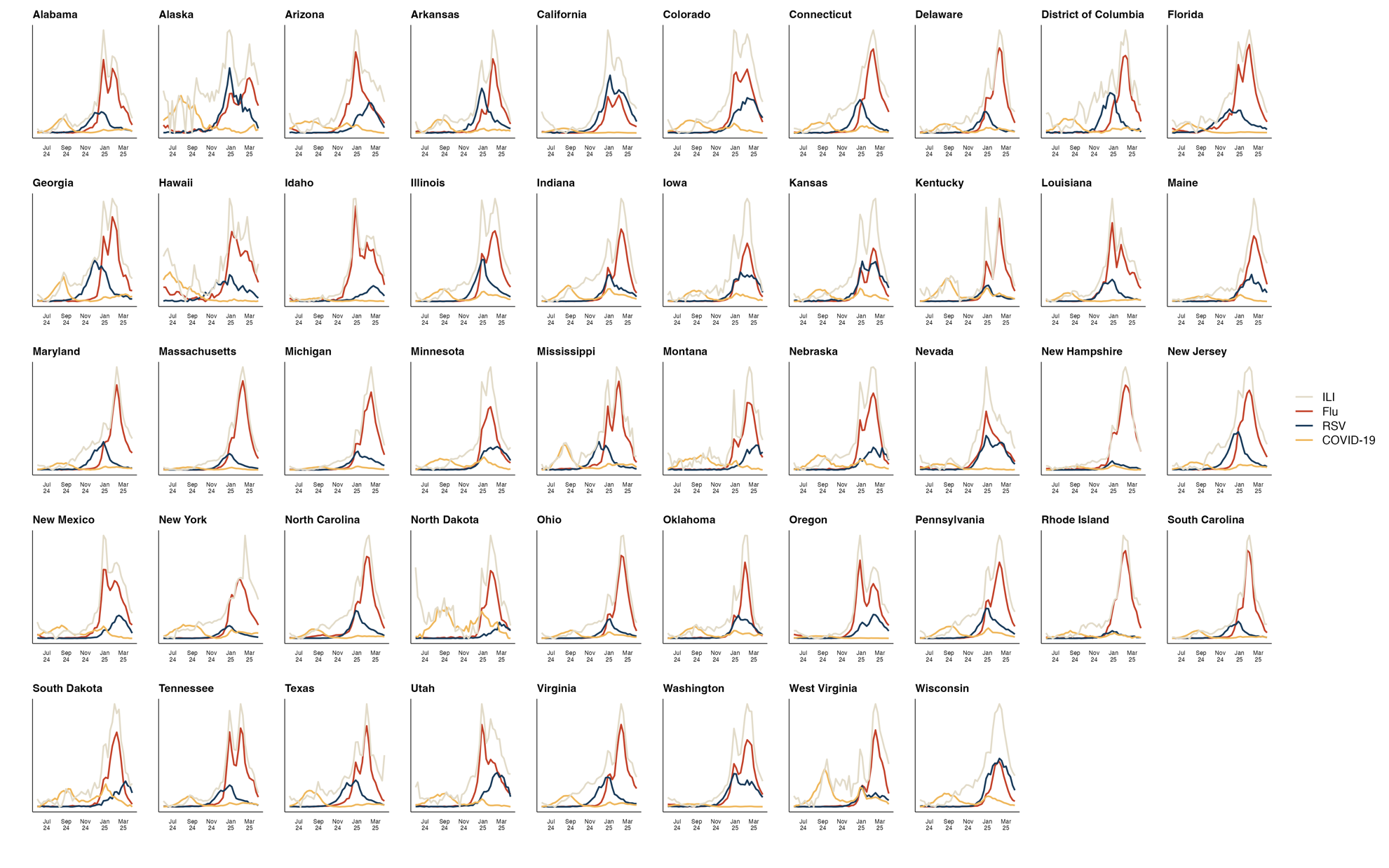

**Supplementary Figure 3: Timing of Respiratory Epidemics in the United States for the 2024-2025 ILI season.** Data from Wyoming and Vermont were not supplied to either NSSP or ILINet and were excluded from the analysis. Abbreviations - COVID-19: coronavirus disease 2019; Flu: influenza; ILI: influenza-like illness; RSV: respiratory syncytial virus

**Lagged Correlation Analysis**

To quantify the temporal relationship between epidemics of influenza, RSV, and COVID-19 and confirm what we observed in our regression modeling results, we deployed a lagged correlation analysis as an alternative to our early warning system method. We first evaluated the time differences between peaks of epidemic activity by computing correlations between the trend of influenza ED visits and lagged trends of both RSV and COVID-19 ED visits (**Supplementary Figures 4-6**). We found that the influenza signal was highly correlated (Pearson correlation ≥ 0.6) with either concurrent RSV, lag-1 RSV, or lag-2 RSV (RSV shifted one or two weeks prior) in 49 out of 50 regions (98.0%) in the 22-23 season, 46 out of 50 regions (92.0%) in the 23-24 season, and 41 out of 48 regions (85.4%) in the 24-25 season. Based on this finding, we estimate that there is up to a 3 week delay between peak RSV volume and peak influenza volume. To validate these findings, we next performed a robustness check by conducting a lagged correlation analysis comparing a fixed influenza signal and COVID-19 signals shifted forward and backward in time (**Supplementary Figures 7-9**). Results of this check show the same inconsistent seasonality that we observed from the regression modeling results – we observe three different correlation patterns across the 3 analyzed seasons, indicating no regularity in the time difference between peaks of influenza volume and peaks of COVID-19 volume.

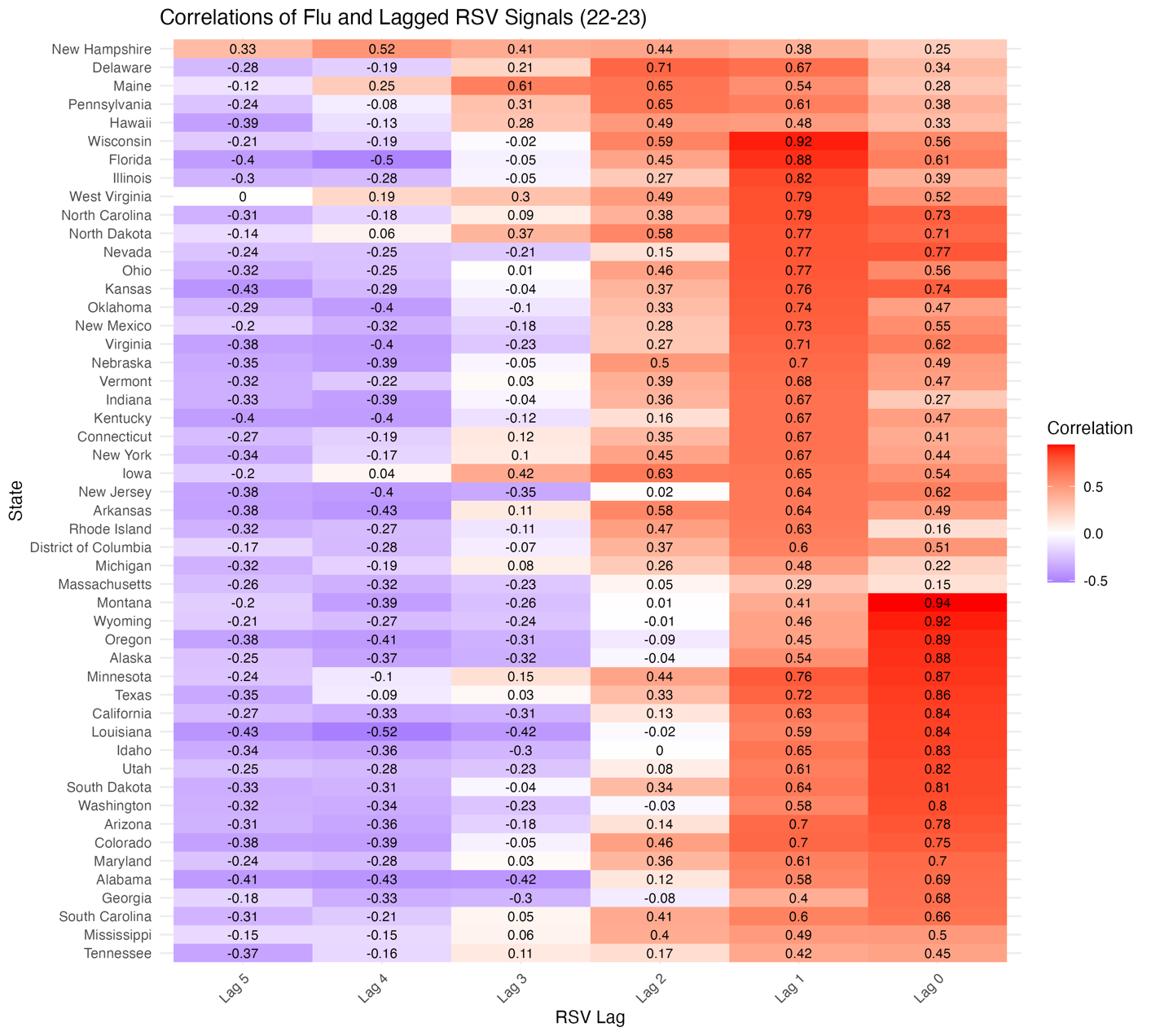

**Supplementary Figure 4: Correlations between influenza ED visits and lagged RSV ED visits in the 2022-23 ILI season.** The strongest correlations between influenza and RSV ED visits in the 22-23 season were in the lag-1 and lag-0 columns, implying the average deviation between signal peaks was between 0 and 1 weeks.

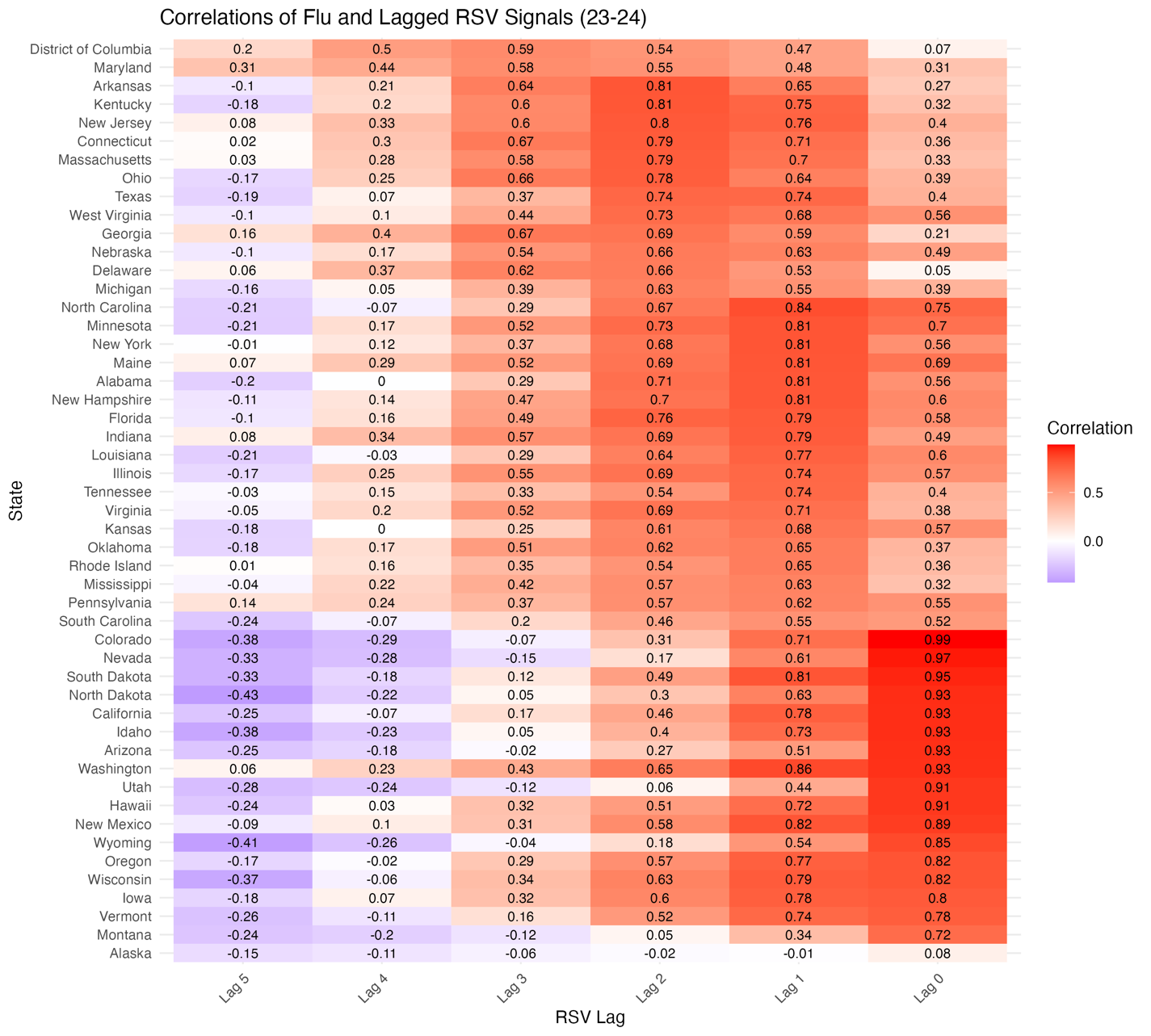

**Supplementary Figure 5: Correlations between influenza ED visits and lagged RSV ED visits in the 2023-24 ILI season.** The strongest correlations between influenza and RSV ED visits in the 23-24 season were in the lag-1 and lag-0 columns, implying the average deviation between signal peaks was between 0 and 1 weeks.

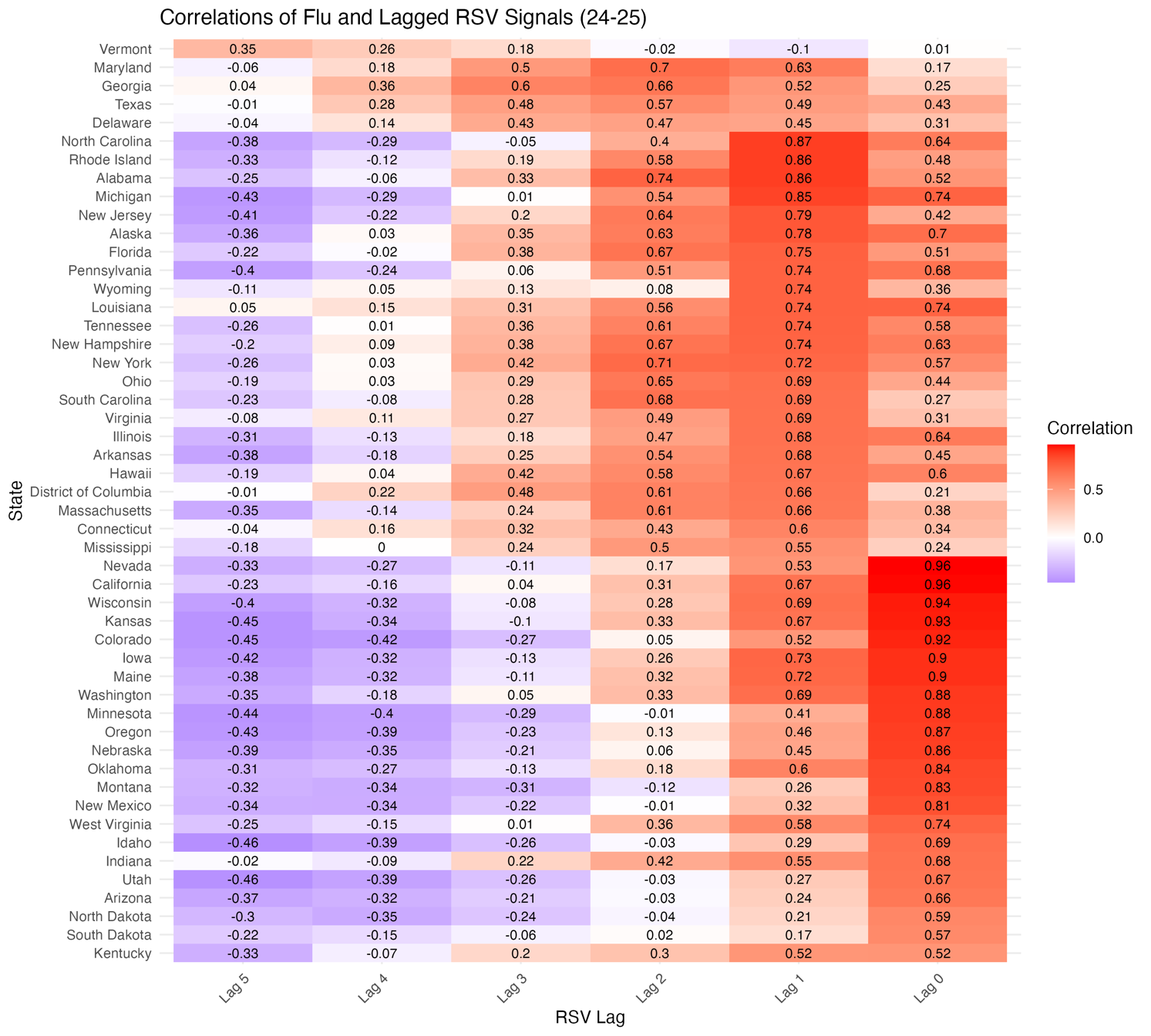

**Supplementary Figure 6: Correlations between influenza ED visits and lagged RSV ED visits in the 2024-25 ILI season.** The strongest correlations between influenza and RSV ED visits in the 24-25 season were in the lag-1 and lag-0 columns, implying the average deviation between signal peaks was between 0 and 1 weeks.

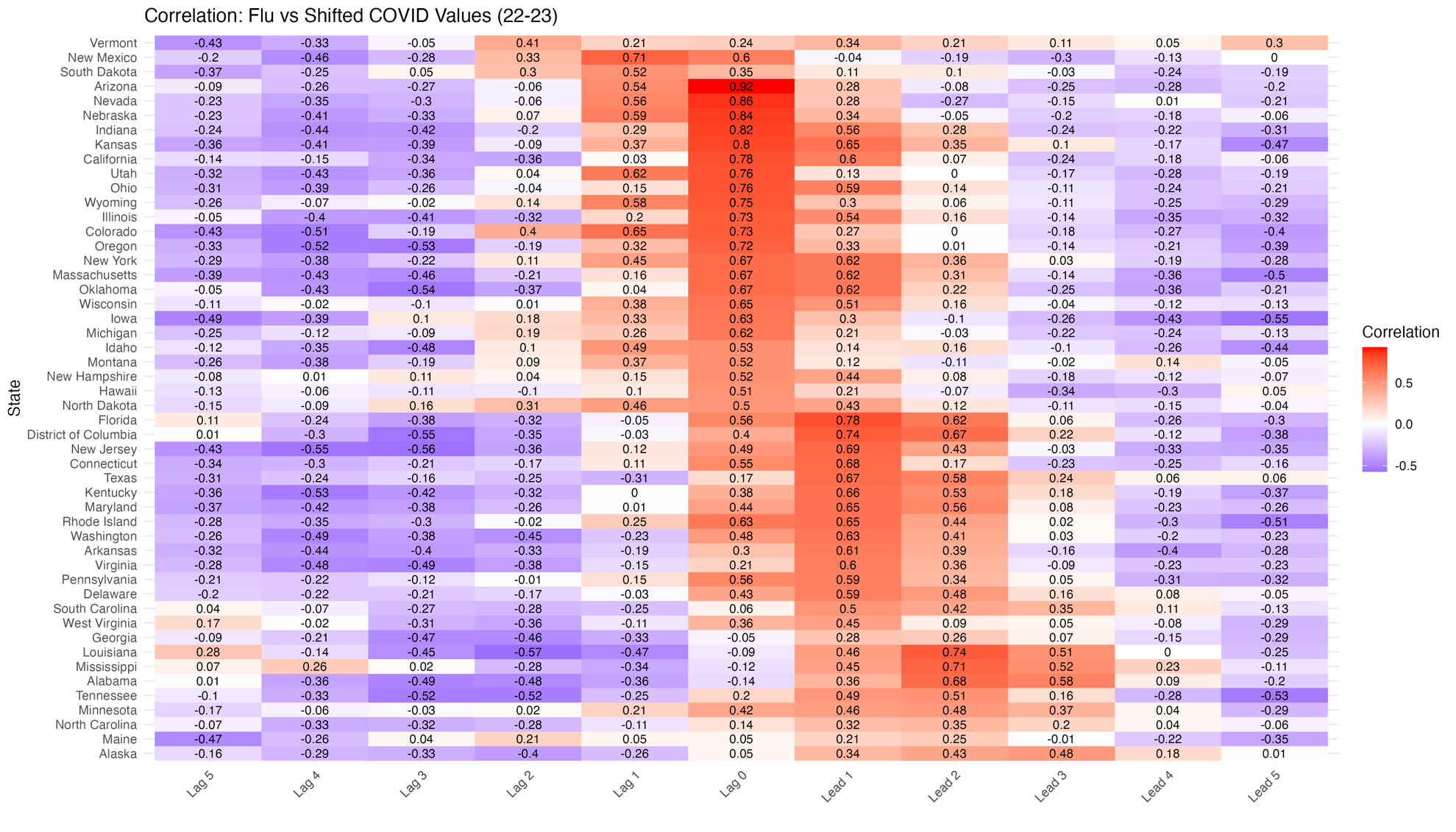

**Supplementary Figure 7: Correlations between influenza ED visits and time-shifted COVID-19 ED visits in the 2022-23 ILI season.**

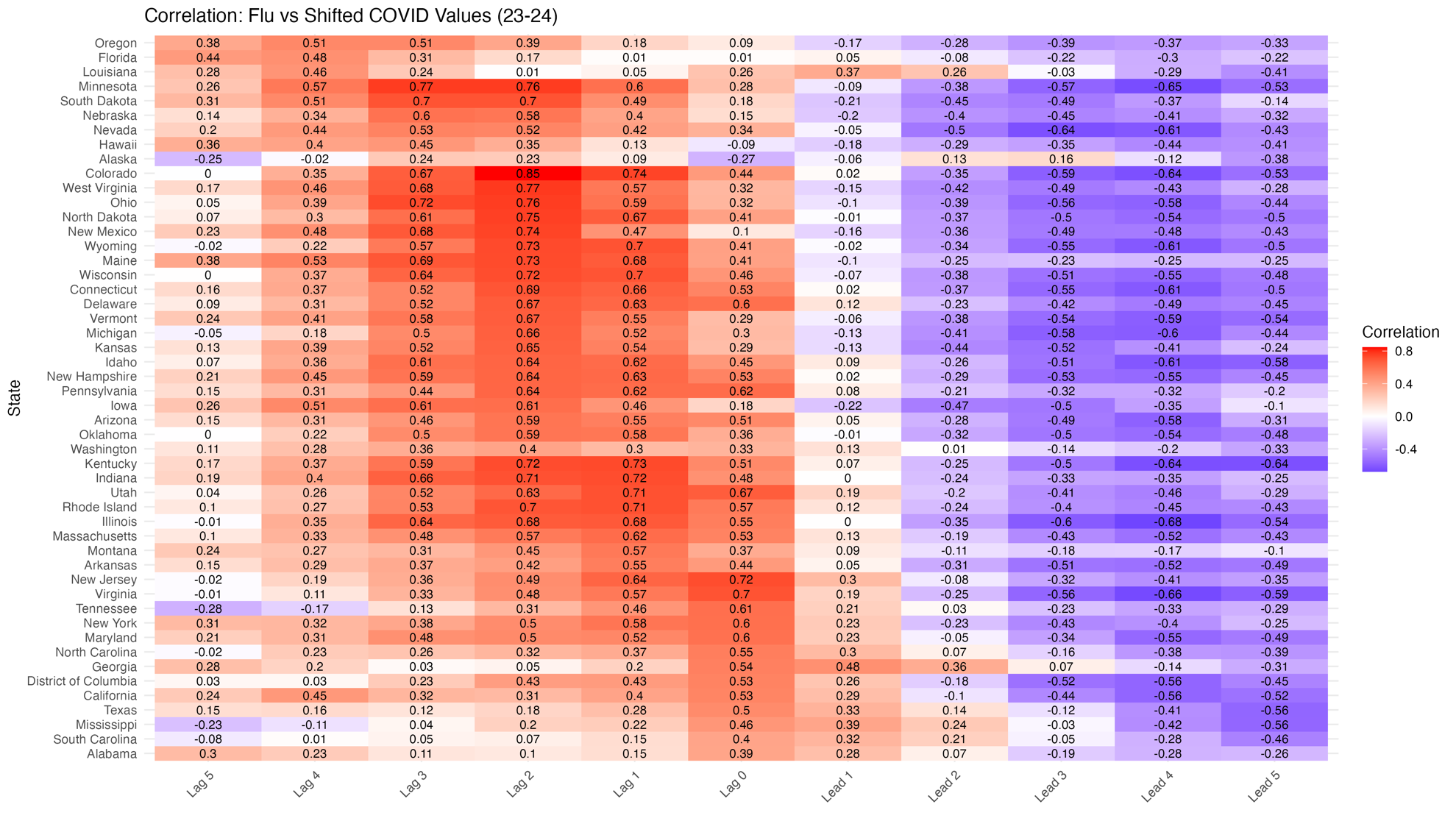

**Supplementary Figure 8: Correlations between influenza ED visits and time-shifted COVID-19 ED visits in the 2023-24 ILI season.**

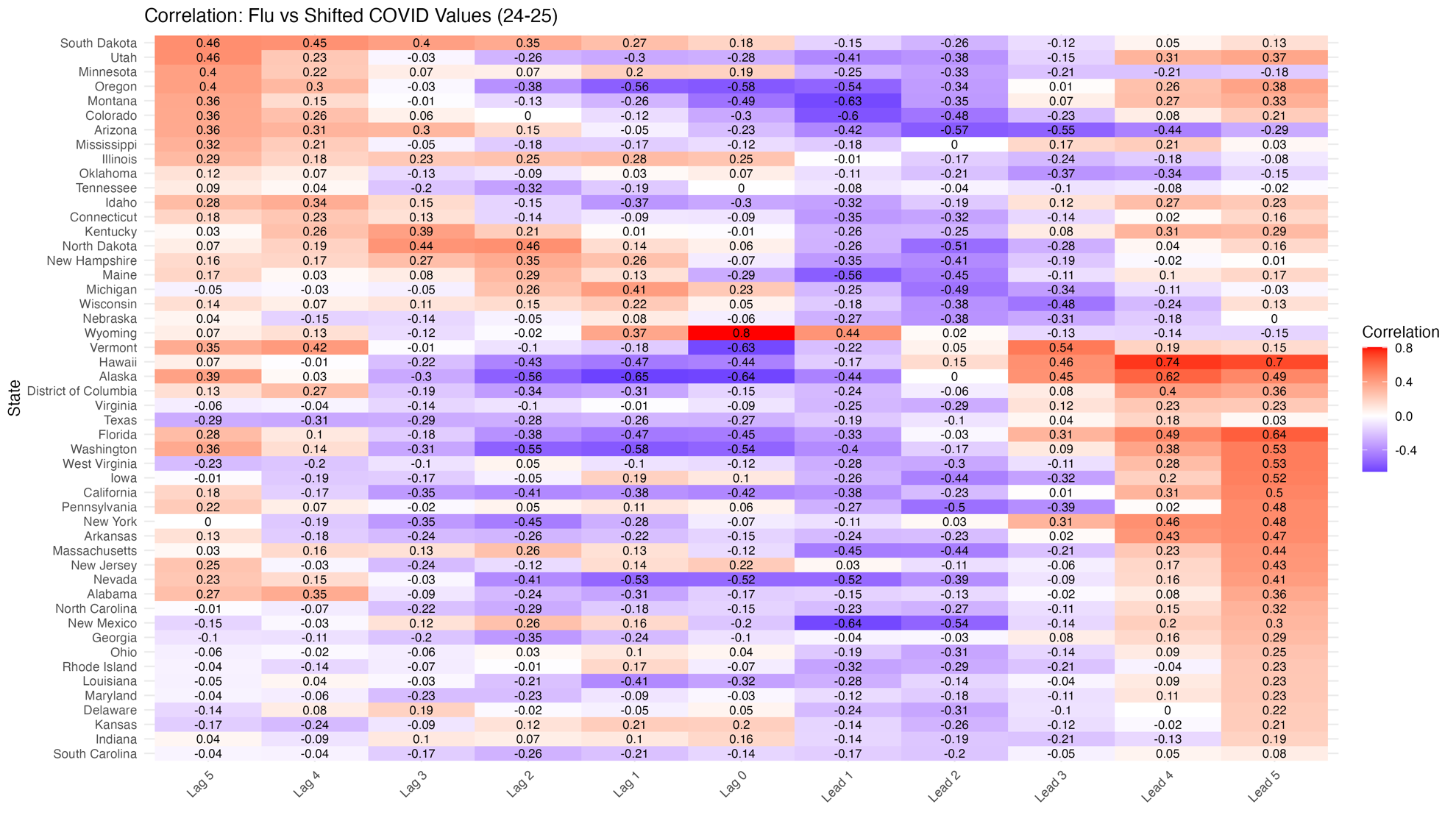

**Supplementary Figure 9: Correlations between influenza ED visits and time-shifted COVID-19 ED visits in the 2024-25 ILI season.**

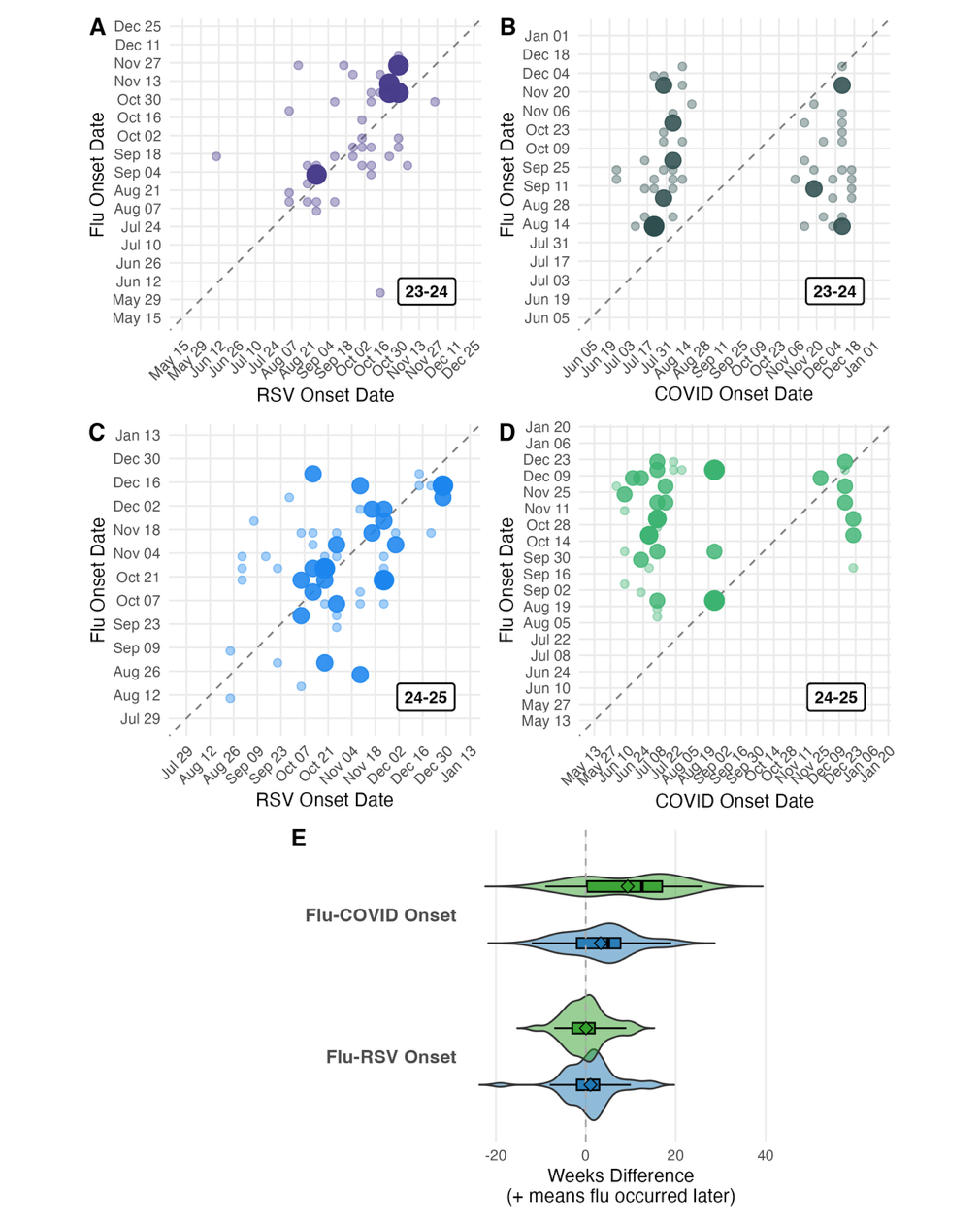

**Supplementary Figure 10: Early warning systems show no clear precedence of RSV or COVID-19 before influenza when comparing epidemic onsets.** Points above the diagonal line indicate that the disease on the *y*-axis (influenza) peaks before the disease on the *x*-axis (either RSV or COVID-19). Larger points indicate that peaks in multiple states occurred during the same week.

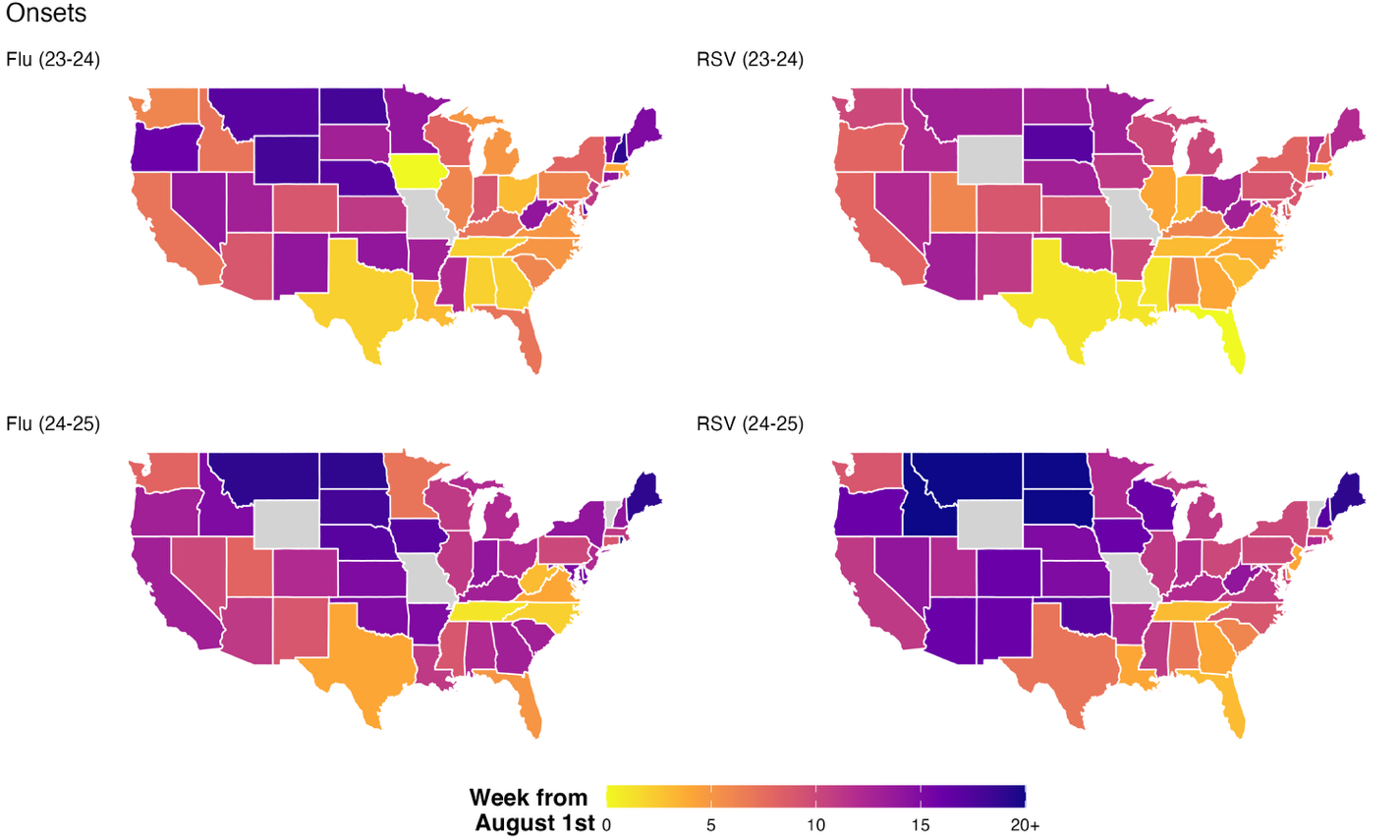

**Supplementary Figure 11: Early warning systems indicate influenza and RSV epidemic onsets spread in a southeast to northwest direction the 2023-24 and 2024-25 ILI seasons.** The colors represent the number of weeks after August 1^st^ of the respective season for the early warning system to detect onset of an epidemic. States that are greyed out did not report data during the corresponding seasons.

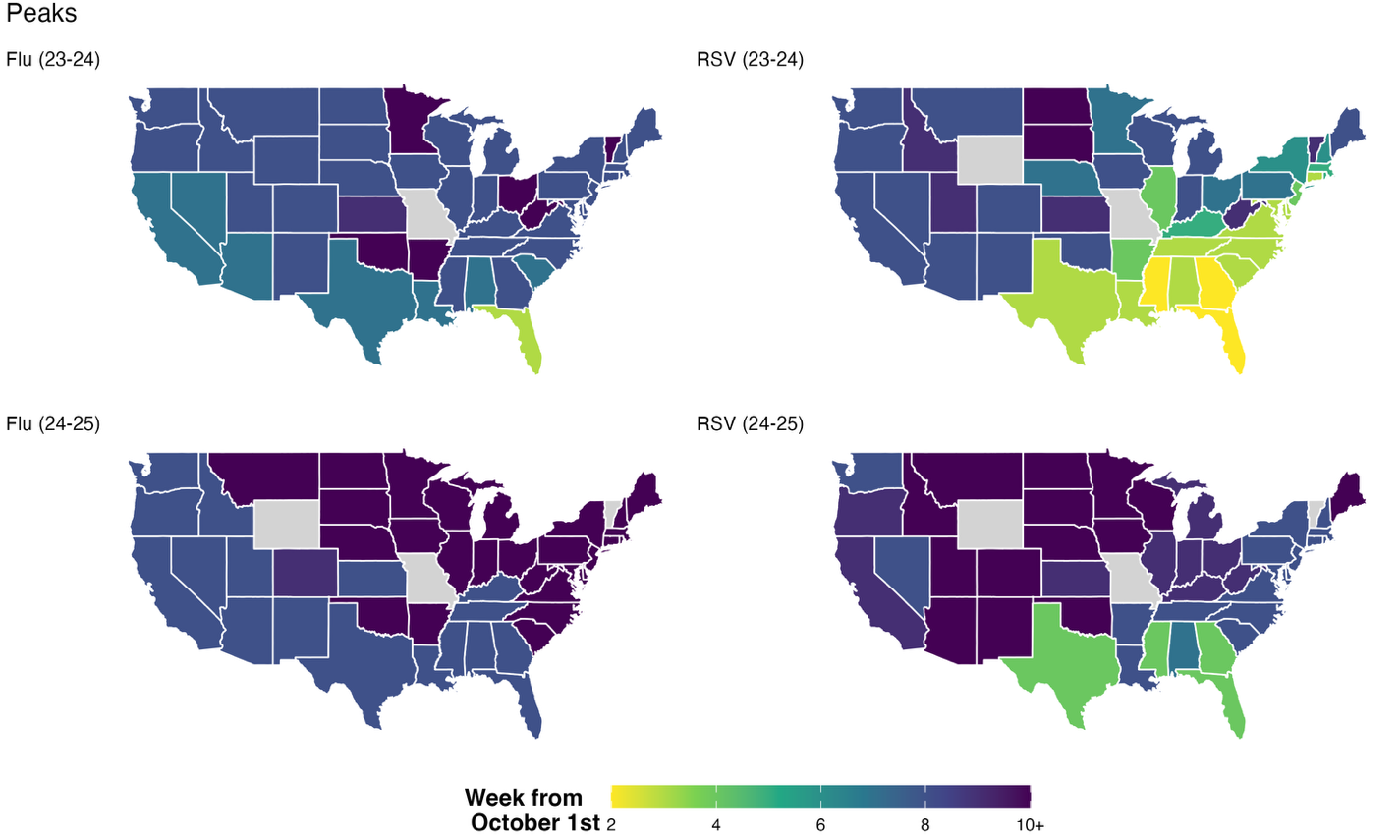

**Supplementary Figure 12: Early warning systems indicate influenza and RSV epidemic peak spread in a southeast to northwest direction the 2023-24 and 2024-25 ILI seasons.** The colors represent the number of weeks after October 1^st^ of the respective seasons for the early warning system to detect an epidemic peak. States that are greyed out did not report data during the corresponding seasons.
